# Development of Diagnostic Score for Acute Pericarditis in Patients Admitted for Chest Pain

**DOI:** 10.1101/2020.08.19.20176750

**Authors:** André Costa Meireles, João Vitor Miranda Porto Oliveira, Bruna de Sá Barreto Pontes, Alexandre Costa Souza, Laila Caroline Oliveira Souza Barbosa Gomes, Thomaz Emanoel Azevedo Silva, Milton Henrique Vitoria de Melo, Gabriela Oliveira Bagano, Márcia Maria Noya Rabelo, Luis Claudio Lemos Correia

## Abstract

**Background:** Despite the presence of clinical, laboratory and electrocardiographic characteristics suggestive of acute pericarditis, there is no multivariate diagnostic score developed for this condition.

**Objective:** To develop a clinical score for diagnosis of pericarditis as the cause of acute chest pain, using data from admission.

**Methods:** In a diagnostic case-control study, we compared consecutive 45 patients of the Chest Pain Registry diagnosed of pericarditis (confirmed by magnetic resonance imaging or the presence of pleural effusion in echocardiography) versus 90 patients with an alternative confirmed diagnosis, randomly selected from our registry. Six clinical characteristics, 16 chest pain characteristics and 4 additional tests were candidates as predictors. Logistic regression was used to derivate a model composed by independent predictors of pericarditis.

**Results:** Among 17 variables associated with pericarditis, 5 remained independent predictors: age; pain aggravation with thorax movement; positive troponin; diffuse ST-segment elevation and C-reactive protein. Each independent predictor was attributed points proportional to its regression coefficient. The final score presented discriminatory capacity represented by C-statistic of 0.97 (95% CI = 0.93 – 1.0). The best cutoff point was defined as > 6 points, with sensitivity of 96% (95% CI = 85 – 100), specificity of 87% (95% CI = 78 – 93), positive likelihood ratio of 7.2 (95% CI = 4.2 – 12) and negative likelihood ratio of 0.05 (95% CI = 0.01 – 0.2).

**Conclusion:** The proposed multivariate score seems to be accurate for the diagnosis of pericarditis and require further validation in an independent sample.

## INTRODUCTION

Acute pericarditis is a frequent cause of chest pain (1,2), mostly representing low risk conditions with expectant treatment, unlike other causes such as acute coronary syndromes, pulmonary embolism, pneumonia and aortic dissection (3-5). Therefore, a differential diagnosis of pericarditis in the acute chest pain scenario has prognostic and treatment implications. However, there is no definitive diagnostic test for acute pericarditis (6,7).

It has been proposed that the numbers of suggestive characteristics define the diagnosis (11), including pericardial friction, electrocardiogram and imaging methods (8-10). However, there is no algorithm derived from epidemiological data and validated in its accuracy. A diagnostic process based on multivariate scores allows the simultaneous use of a set of variables, structuring a value hierarchy for each variable. This discriminatory process tends to be more accurate than the disorderly influence of medical thinking based on clinical impression (12,13). To our knowledge, there are no validated probabilistic models for diagnosing pericarditis.

Thus, in the present study of diagnostic case-control, we identified independent predictors and derived a multivariate score to discriminate pericarditis in a random set of other etiological conditions.

## METHODS

### Sample Selection

During the period from 2014 to 2018, all patients admitted due to chest pain in the coronary unit of a tertiary hospital were included in a prospective data record. From this record, it was selected all 45 patients with a magnetic resonance imaging positive for pericarditis and 90 control patients with a proven alternative etiology, randomly selected.

Pericarditis was defined by magnetic resonance demonstrating pericardial edema, pericardial enhancement, myocardial enhancement, non-subendocardial myocardial enhancement or pericardial effusion or echocardiogram showing pericardial effusion. Controls were defined by the following proven diagnosis: obstructive coronary disease (stenosis ≥ 70% by invasive coronarography); pulmonary embolism (confirmed by chest angio-CT); pneumonia (infectious condition and confirmation by a radiological exam); aortic dissection (confirmed by chest angio-CT); or gastrointestinal disease (ulcer or high grade inflammation on upper digestive endoscopy). The random selection was made by the patient’s number on the registry.

The only exclusion criterion was the patient’s refusal to participate the study. The protocol is in accordance with the Declaration of Helsinki, approved by Institution’s Research Ethics Committee and all patients signed the informed consent form.

### Predictor Variables

On admission, three groups of variables were registered as candidates to pericarditis prediction: medical history, pain characteristics and complementary exams. The first group included age, sex and previous history of smoking, systemic arterial hypertension, diabetes mellitus and coronary artery disease (CAD). The second group was composed by 16 chest pain characteristics; and the third set of variables included widespread ST segment elevation on electrocardiogram (≥ 1 mm in more than one electrocardiographic wall), depressed PR interval (≥ 1 mm in more than one lead), positive troponin (Ortho-Clinical Diagnostics, Rochester, NY, EUA) and numeric value of high sensitivity C-reactive protein (nephelometry, Dade-Behring, EUA). Positive troponin was defined as any value above detection limit (> 0,012 ug/L), not using 99 percentile because it is the definition of myocardial infarction (14). Laboratory tests were performed on plasma collected at presentation in the emergency room. Medical history and chest pain characteristics were recorded by researchers trained to interview patients systematically, in order to reduce bias and improve reproducibility. Patients’ electrocardiogram was interpreted blindly by two trained cardiologists (they were unaware of patient’s history and diagnosis). The disagreement of any of the reports was discussed between the two cardiologists until a consensus was reached.

### Statistical Analysis

Categorical variables were described by frequency, while numerical variables of normal distribution were expressed by mean and standard deviation, and numerical variables of non-normal distribution by median and interquartile range. The program used for statistical analysis was SPSS (Version 25, SPSS Inc., Chicago, Illinois, USA).

Statistical analysis is represented in Figure 1. First, it was tested univariate associations between pericarditis and 26 variables candidates to predictors by Pearson’s chi-square for categorical variables and unpaired t student’s *t* test for numerical variables. Variables associated with pericarditis with a P value < 0.10 were included in the multivariate logistic regression analysis. In order to reduce the number of predictors in the final logistic analysis, intermediate logistic regression models were used for variables related to medical history and variables related to pain characteristics. Significant variables (P < 0.10) from the intermediate models and complementary tests that were associated with the outcome in univariate analysis entered the final analysis. Due to the premise that age influence on the diagnosis of pericarditis is not linear, for the final logistic regression model, age was categorized in four pre-defined groups by decades. Statistical significance in the final model was defined as P < 0.05.

**Figure 1:**
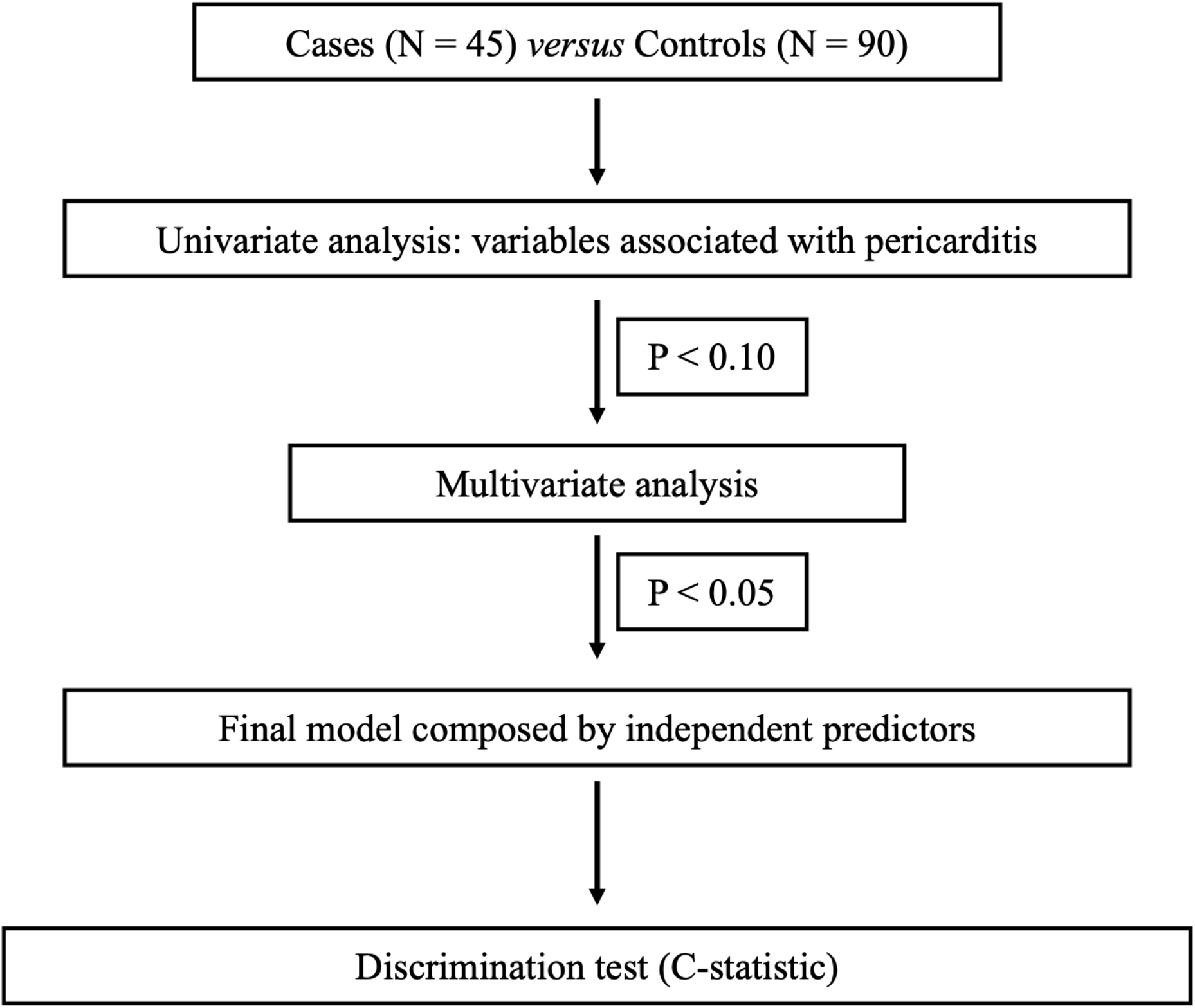
Flow chart of statistical analysis.

For the creation of a point-based score, regression coefficients were approximated to the nearest whole value, which consisted of the points of score to each positive variable. In calculating the score, missing values were imputed by the most frequent in categorical variables and by the mean value in numerical variables. The score of each patient was assessed in relation to their discriminatory capacity by the area below the ROC curve. Comparison of the score between the two groups was performed using Mann-Whitney non-parametric test

## RESULTS

### Sample Characteristics

The pericarditis group was composed by 45 patients, 87% males, age 36 ± 14 years old (median 34, interquartile range 25 to 43, absolute amplitude 16 to 70). The control group consisted of 90 patients, distributed in 41 diagnosis of myocardial infarction, 29 unstable angina, 10 pulmonary embolisms, 2 aortic dissections, 2 pneumonias and 2 with gastrointestinal pain – Figure 2. The mean age of the control group (61 ± 15 anos) was significantly higher than patients with pericarditis (P < 0.001), with no male dominance (57%) as observed in the pericarditis group – Table 1.

**Figure 2:**
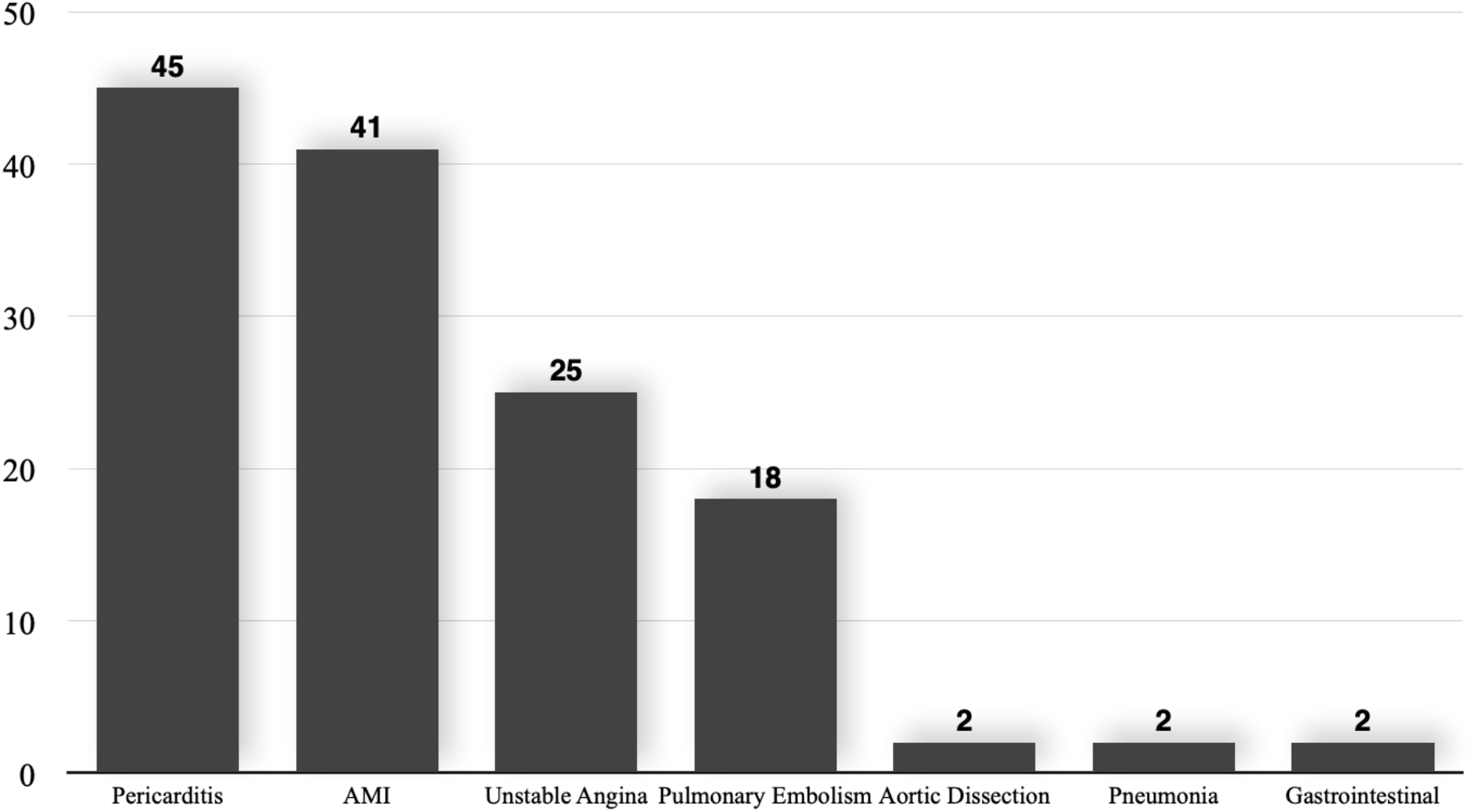
Absolute diagnostic frequency of the sample, showing a greater predominance of acute coronary syndrome in the control group, followed by pulmonary embolism

**Figure 3:**
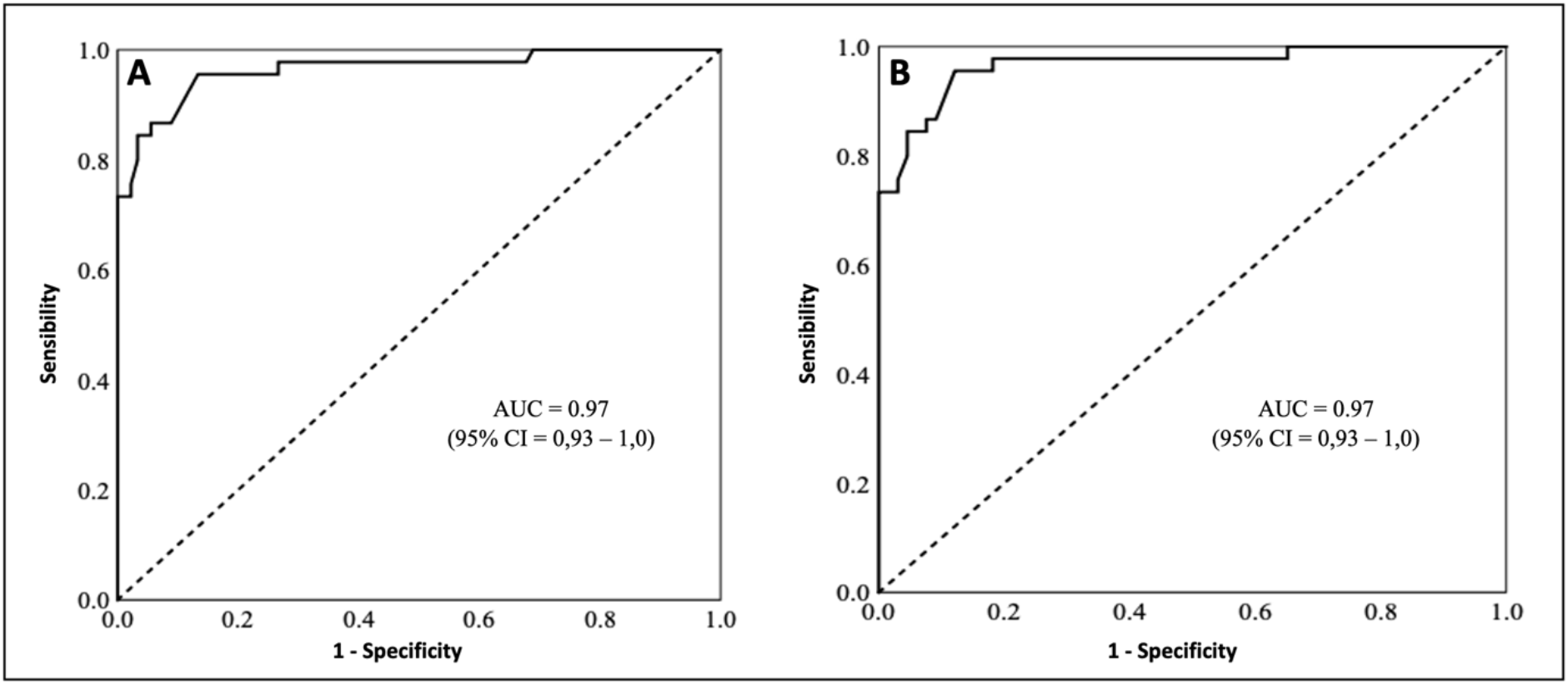
Panel A shows ROC curve of the predictor model discriminating between pericarditis and other causes. Panel B shows the ROC curve of discrimination between pericarditis and acute coronary syndrome.

**Table 1.**
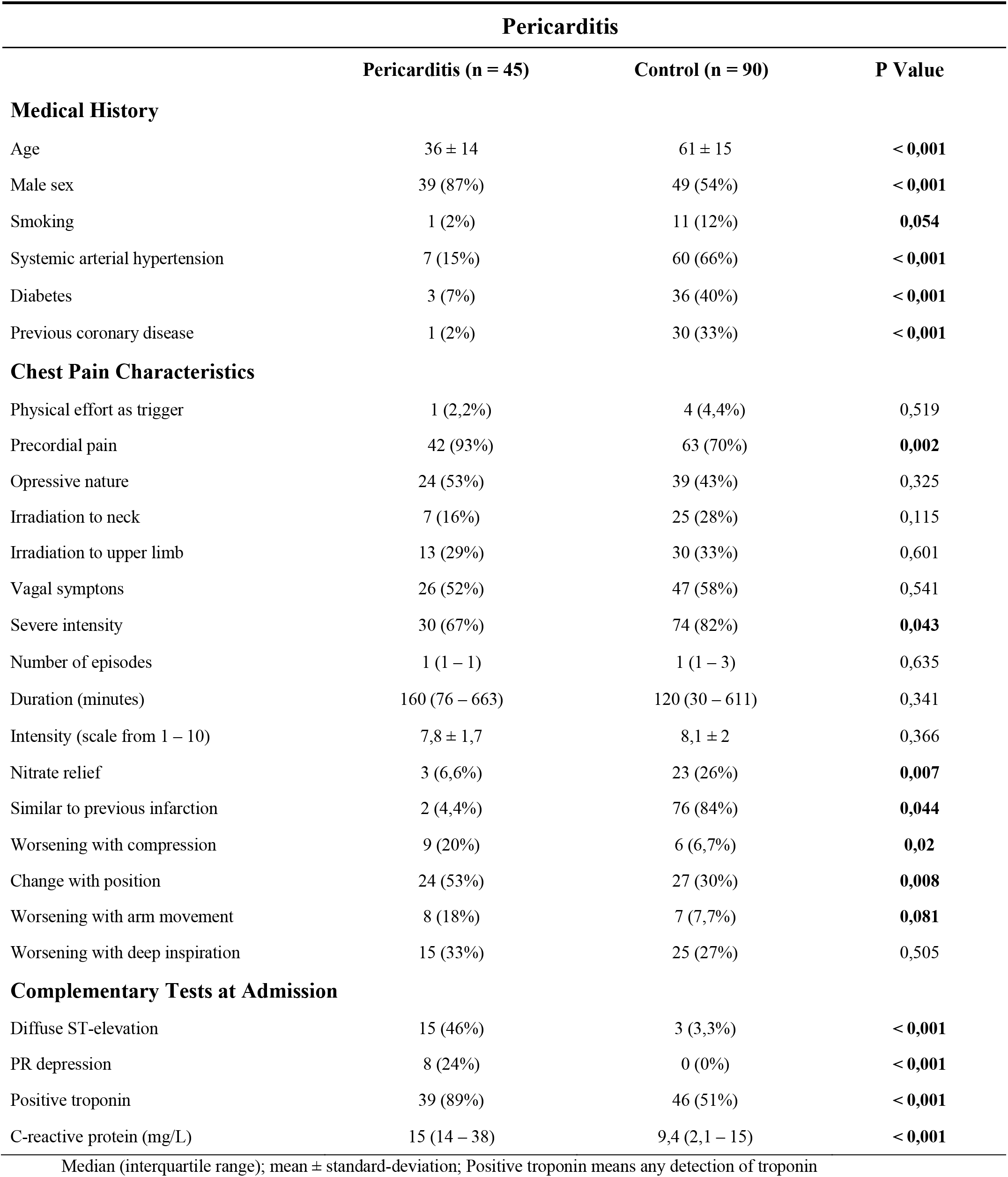
Comparison of medical history, chest pain characteristics and complementary tests between patients diagnosed with pericarditis and patients with another established diagnosis.

| **Pericarditis** |  |  |  |
| --- | --- | --- | --- |
|  | **Pericarditis (n = 45)** | **Control (n = 90)** | **P Value** |
| **Medical History** |  |  |  |
| Age | 36 ± 14 | 61 ± 15 | < 0,001 |
| Male sex | 39 (87%) | 49 (54%) | < 0,001 |
| Smoking | 1 (2%) | 11 (12%) | 0,054 |
| Systemic arterial hypertension | 7 (15%) | 60 (66%) | < 0,001 |
| Diabetes | 3 (7%) | 36 (40%) | < 0,001 |
| Previous coronary disease | 1 (2%) | 30 (33%) | < 0,001 |
| **Chest Pain Characteristics** |  |  |  |
| Physical effort as trigger | 1 (2,2%) | 4 (4,4%) | 0,519 |
| Precordial pain | 42 (93%) | 63 (70%) | 0,002 |
| Opressive nature | 24 (53%) | 39 (43%) | 0,325 |
| Irradiation to neck | 7 (16%) | 25 (28%) | 0,115 |
| Irradiation to upper limb | 13 (29%) | 30 (33%) | 0,601 |
| Vagal symptoms | 26 (52%) | 47 (58%) | 0,541 |
| Severe intensity | 30 (67%) | 74 (82%) | 0,043 |
| Number of episodes | 1 (1 – 1) | 1 (1 – 3) | 0,635 |
| Duration (minutes) | 160 (76 – 663) | 120 (30 – 611) | 0,341 |
| Intensity (scale from 1 – 10) | 7,8 ± 1,7 | 8,1 ± 2 | 0,366 |
| Nitrate relief | 3 (6,6%) | 23 (26%) | 0,007 |
| Similar to previous infarction | 2 (4,4%) | 76 (84%) | 0,044 |
| Worsening with compression | 9 (20%) | 6 (6,7%) | 0,02 |
| Change with position | 24 (53%) | 27 (30%) | 0,008 |
| Worsening with arm movement | 8 (18%) | 7 (7,7%) | 0,081 |
| Worsening with deep inspiration | 15 (33%) | 25 (27%) | 0,505 |
| **Complementary Tests at Admission** |  |  |  |
| Diffuse ST-elevation | 15 (46%) | 3 (3,3%) | < 0,001 |
| PR depression | 8 (24%) | 0 (0%) | < 0,001 |
| Positive troponin | 39 (89%) | 46 (51%) | < 0,001 |
| C-reactive protein (mg/L) | 15 (14 – 38) | 9,4 (2,1 – 15) | < 0,001 |

Median (interquartile range); mean ± standard-deviation; Positive troponin means any detection of troponin

### Intermediate Predictors of Pericarditis

All variables related to medical history showed statistically significant difference between the pericarditis group and the non-pericarditis group: age, male sex, smoking, hypertension, diabetes and previous coronary disease – Table 1. The frequency of pericarditis in the age groups < 30 years old, 30-39 years old, 40-49 years old and > 50 years old was 95%, 59%, 44% and 10%, respectively (P < 0.001). When these variables were included in the logistic regression model, age, male sex and previous CAD remained statistically significant (intermediate Model 1 – Table 2).

As for the 16 chest pain characteristics, seven showed significant association with pericarditis: precordial pain, severe intensity, nitrate relief, similarity with previous acute myocardial infarction, worsening with compression, worsening with change in position and worsening with arm movement – Table 1. Of these variables, precordial pain, nitrate relief and worsening with change in position were the three independent predictors in intermediate Model 2 – Table 2.

**Table 2.**
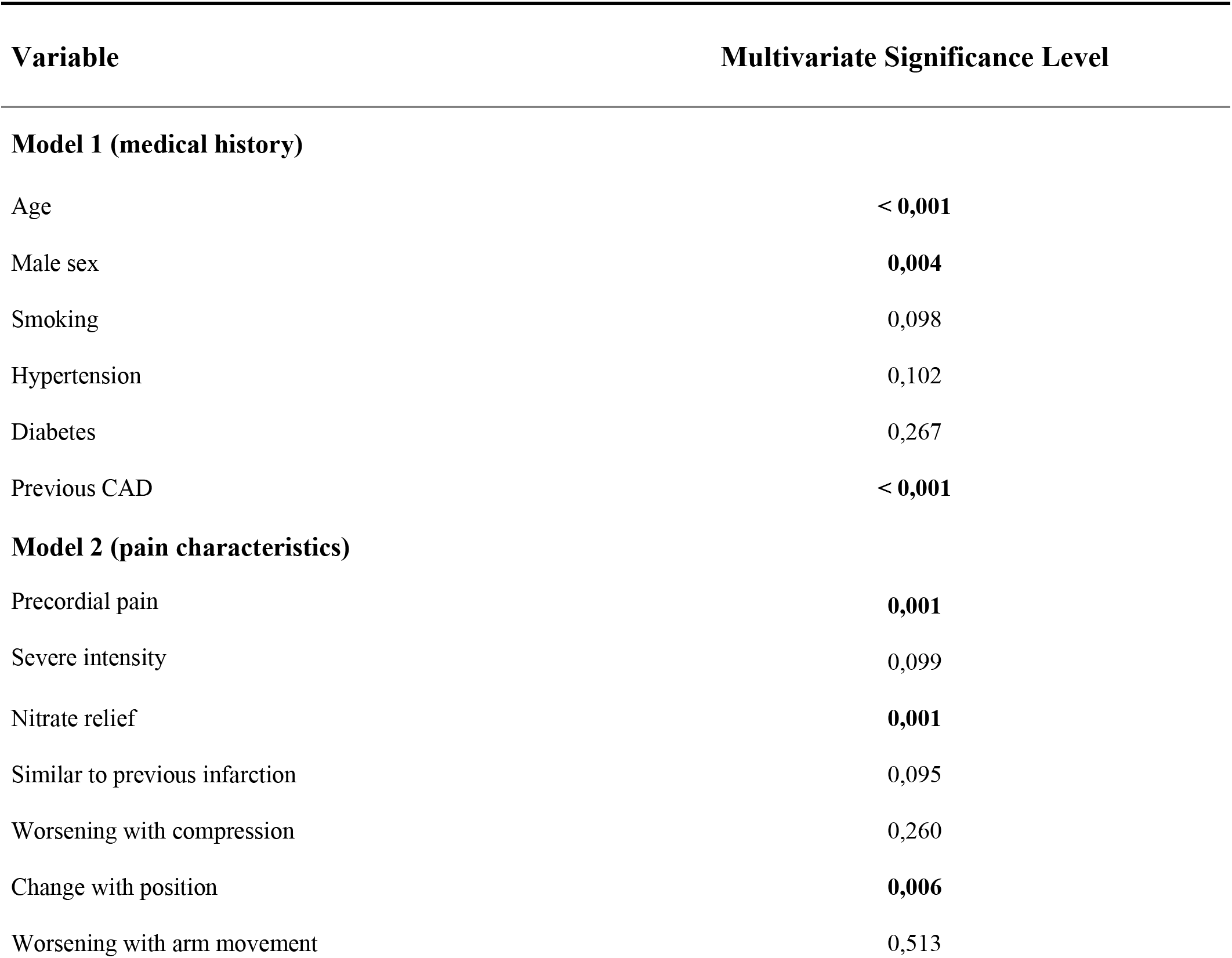
– Intermediate models of logistic regression of medical history (Model 1) and characteristics of chest pain (Model 2).

| Variable | Multivariate Significance Level |
| --- | --- |
| **Model 1 (medical history)** |  |
| Age | < 0,001 |
| Male sex | 0,004 |
| Smoking | 0,098 |
| Hypertension | 0,102 |
| Diabetes | 0,267 |
| Previous CAD | < 0,001 |
| **Model 2 (pain characteristics)** |  |
| Precordial pain | 0,001 |
| Severe intensity | 0,099 |
| Nitrate relief | 0,001 |
| Similar to previous infarction | 0,095 |
| Worsening with compression | 0,260 |
| Change with position | 0,006 |
| Worsening with arm movement | 0,513 |

Regarding complementary exams, all showed significant difference in univariate analysis: diffuse ST elevation, PR depression, positive troponin and the value of C-reactive protein – Table 1. These variables were directly included in the final regression analysis.

### Independent Predictors of Pericarditis

The 13 variables independently associated with pericarditis in intermediate models 1 and 2 and the four variables related to complementary tests were candidates to the final analysis, which was built from a logistic regression using the stepwise method.

Considering the predictive value of age better related to younger age groups, for the construction of the score, age was categorized a priori as < 30 years old, 30-39 years old, 40-49 years old and > 50 years old as the reference age range.

Independent predictors of pericarditis in the multivariate analysis were: age < 30 years old (OR = 141; 95% CI = 1.9 – 9464); 30-39 years old (OR = 18; 95% CI = 2.6 – 100); 40-49 years old (OR = 3.6; 95% CI = 0.32 – 27)]; change in pain with position (OR = 6.8; 95% CI = 2.1 – 54); positive troponin (OR = 41; 95% CI = 4.7 – 448); diffuse ST elevation (OR = 74; IC 95% = 1.3 – 4201) e C-reactive protein (OR = 1.014; 95% IC = 1.001 – 1.026) – Table 3.

**Table 3.** – Final logistic regression model defining independent predictors of pericarditis.

| Variable | βeta | Odds Ratio (95% CI) | P Value |
| --- | --- | --- | --- |
| Age (years old) |  |  |  |
| < 30 | 5,5 | 254 (4 – 16142) | 0,009 |
| 30 - 39 | 3,0 | 21 (3,6 – 124) | 0,001 |
| 40 - 49 | 1,6 | 5,0 (0,6 – 39) | 0,125 |
| > 50 | Reference |  |  |
| Change in pain with position | 2,4 | 11 (2,0 – 54) | 0,004 |
| Diffuse ST elevation | 3,8 | 43 (3,8 – 484) | 0,022 |
| Positive troponin | 3,8 | 47 (5,0 – 437) | 0,001 |
| C-reactive protein | 0,015 | 1,015 (1,003 – 1,03) | 0,017 |
| *Constant* | -7,388 | ---- | ---- |
| **Excluded Variables** |  |  |  |
| Male sex | ---- | ---- | 0,193 |
| Previous CAD | ---- | ---- | 0,871 |
| Precordial pain | ---- | ---- | 0,941 |
| Nitrate relief | ---- | ---- | 0,174 |
| PR depression | ---- | ---- | 0,999 |

When these predictors were analyzed individually, the area under the curve of age was 0.87 (95% CI = 0.81 – 0.94) and of C-reactive protein 0.76 (95% CI = 0.68 – 0.84) – Table 4. Sensibility, specificity and positive and negative likelihood ratio of the variables change in pain with position, positive troponin and widespread ST elevation are shown in Table 4.

**Tabela 4.** – Accuracy of isolated variables that composed the final predictor model.

| Dichotomous Variable | Sensibility | Specificity | Positive Likelihood Ratio | Negative Likelihood Ratio |
| --- | --- | --- | --- | --- |
| Change in pain with position | 53%<br>(95% CI = 38 - 68) | 70%<br>(95% CI = 59 - 79) | 1,8<br>(95% CI = 1,2 - 2,7) | 0,67<br>(95% CI = 0,47 - 0,94) |
| Diffuse ST elevation | 45%<br>(95% CI = 28 - 64) | 97%<br>(95% CI = 91 - 99) | 13<br>(95% CI = 4 - 44) | 0,56<br>(95% CI = 0,41 - 0,77) |
| Positive troponin | 89%<br>(95% CI = 75 - 96) | 49%<br>(95% CI = 38 - 60) | 1,63<br>(95% CI = 1,4 - 2,2) | 0,23<br>(95% CI = 0,10 - 0,54) |
| Numerical Variable | Area Under ROC |  | Confidence Interval (95%) |  |
| Age | 0,87 |  | 0,81 - 0,94 |  |
| C-reactive protein | 0,76 |  | 0,68 - 0,84 |  |

### Pericarditis Predictor Score

Following proportionality with regression coefficients, each variable received different points for the formation of the score: widespread ST elevation: 4 points; positive troponin: 4 points; change in pain with position: 2 points; C-reactive protein: numerical value of C-reactive protein value multiplied by 0.014. Age range < 30 years old, 30-39 years old, 40-49 years old and > 50 years old respectively received 5 points, 3 points, 1 point and 0 points – Table 5.

**Table 5.** – Final predictor score and its respective points

| Variable | Score |
| --- | --- |
| Age (years) |  |
| < 30 | 5 |
| 30 - 39 | 3 |
| 40 - 49 | 1 |
| > 50 | 0 |
| Change in pain with position | 2 |
| Diffuse ST elevation | 4 |
| Positive troponin | 4 |
| C-reactive protein | value x 0.014 |
| **Interpretation** | **Score** |
| Suggestive of Pericarditis | > 6 |
| Suggestive of non-Pericarditis | ≤ 6 |

N: numerical value of C-reactive protein.

The median score of patients with pericarditis was 11 (7.6 – 13), compared to 4.1 (1.0 – 5.2) in the control group (P < 0.001). The score showed excellent discriminatory capacity, with area under the ROC curve of 0.97 (95% CI = 0.93 – 1.0) – Figure 4A. The best cut-off point was > 6 points, with a sensitivity of 96% (95% CI = 885 – 100), specificity of 87% (95% CI = 78 – 93), positive likelihood ratio of 7.2 (95% CI = 4.2 – 12) and negative likelihood ratio of 0. 05 (95% CI = 0.01 – 0.2).

In the distinction between pericarditis and the 70 patients with acute coronary syndromes, the model also presented an area under the ROC curve of 0.97 (95% CI = 0.93 – 1.0) – Figure 4B.

## DISCUSSION

The present study originally proposes a diagnostic score for pericarditis in patients with acute chest pain. The model was extracted from data present in the setting of acute chest pain, adding C-reactive protein as an inflammatory marker. Of the 26 candidate variables, five were consolidated as independent predictors: age, pain variation with body movement, diffuse ST-segment elevation, positive troponin and C-reactive protein. Evaluation carried out in the same sample suggests an excellent accuracy to discriminate pericarditis from other causes, although this finding requires final validation in an independent sample.

In the absence of a gold standard test, the diagnosis of pericarditis is based on the exclusion of other causes and presence of suggestive clinical data (syndromic diagnosis). Paradoxically to the multifactorial nature of the diagnosis, the need to use diagnostic scores in this condition has not been emphasized. For example, the European Guidelines of Pericarditis suggest a simplistic criteria of the presence of two out of four positive criteria: (1) chest pain, typically pleuritic and that changes with position; (2) pericardial friction; (3) widespread ST elevation or PR depression on EKG; and (4) pericardial effusion (8). However, these diagnostic criteria are not proven to be accurate and the low prevalence of friction and effusion raises doubt about the its performance. Unlike the diagnosis based on the number of criteria, a better approach is the use of multivariate models made of independent predictors weighted according to their level of association.

Our study shows that, in the setting of acute chest pain, clinical and laboratory variables significantly influence the diagnostic probability, demonstrating the potential accuracy of the multivariate and weighted approach (scores). This is a study of conceptual value, which should originate future investigations in order to derive and validate probabilistic models. Although with wide confidence intervals, the point estimate of the five risk predictors suggests that they have a high relative impact on the chance of disease. Among the categorical variables, diffuse ST-elevation is the one with the greatest impact, followed by positive troponin and with less weight, worsening pain with chest movement. It is worth noting that we preferred to treat troponin as a categorical variable, as there is great variation in the measurement methods of this marker. Although troponin is a marker of myocardial injury that increases in many other circumstances, especially infarction, it was able to modulate the likelihood of pericarditis. And the model containing troponin showed good discriminatory capacity between pericarditis and infarction. Among numerical variables, age demonstrated a greater diagnostic influence than inflammation measured by C-reactive protein. In the final model, age was categorized in decades under the premise that its influence is not linear, and its greatest weight is in younger age groups.

The analysis of the PR interval depression and the diffuse characteristic of the ST segment elevation were not predefined characteristics in this prospective record of acute chest pain. For this reason, it was necessary to reanalyze the electrocardiograms, opting for a case-control design, where all cases of pericarditis and a limited number of controls would have the electrocardiogram revised. This analysis was performed blindly in relation to the clinical condition and paired by two independent investigators. This design had some implications: being a case-control study, it was not possible to calculate the prevalence of pericarditis, which limits the calibration of the model to estimate probability. Thus, the present study validates the discriminatory capacity of the score but does not test the calibration of the regression formula. In other words, the tested model appears to be accurate to differentiate pericarditis from non-pericarditis, but not to offer the likelihood of pericarditis. In a future study, we will recalibrate the model across the sample.

On the other hand, the case-control design allows the analysis of the relative value of each covariate. Our model proposes a differentiated view of prediction, not including uncommon findings, such as pericardial friction and pericardial effusion, and based on variables of great impact such as age and electrocardiogram, and new variables in this approach, such as positive troponin and degree of inflammation as C-reactive protein.

In this study, cases of pericarditis were defined by magnetic resonance imaging or pericardial effusion on echocardiogram. Although MRI is the best isolated diagnostic method, its sensitivity and specificity are not perfect (15). This makes us recognize that the inclusion criteria may have left out some typical cases of pericarditis or included false positives. However, it would be inappropriate to involve any other criteria in the definition of cases, as these criteria were being evaluated as candidates for the predictor model.

In conclusion, the present study demonstrates the potential of multivariate and weighted approach in the form of a diagnostic score for the diagnosis of pericarditis in the context of chest pain.

## Data Availability

All data is available

## REFERENCES

1. Lewinter MM. Acute pericarditis. N Engl J Med. 2014;371(25):2410–6.

2. Erhardt L, Herlitz J, Bossaert L, Halinen M, Keltai M, Koster R, et al. Task force on the management of chest pain. Eur Heart J. 2002;23(15):1153–76.

3. Montera MW, Mesquita ET, Colafranceschi AS, de Oliveira AC, Rabischoffsky A, Rochitte CE, et al. I Brazilian guidelines on myocarditis and pericarditis. Arq Bras Cardiol. 2013;100(4 SUPPL):1-36.

4. Imazio M, Gaita F. Diagnosis and treatment of pericarditis. Heart. 2015;101(14):1159-68.

5. Bassan R, Pimenta L, Leães PE, Timerman A. Sociedade Brasileira de Cardiologia: I Diretriz de Dor Torácica na Sala de Emergência. Arq Bras Cardiol. 2002;79(SuplII: 1).

6. Schwefer M, Aschenbach R, Heidemann J, Mey C, Lapp H. Constrictive pericarditis, still a diagnostic challenge: comprehensive review of clinical management. Eur J Cardio-thoracic Surg. 2009;36(3):502–10.

7. Lange RA, Hillis LD. Acute pericarditis. N Engl J Med. 2004;351:2195-202.

8. Adler Y, Charron P. The 2015 ESC Guidelines on the diagnosis and management of pericardial diseases. Eur Heart J. 2015;36(42):2873–4.

9. Klein AL, Abbara S, Agler DA, Appleton CP, Asher CR, Hoit B, et al. American society of echocardiography clinical recommendations for multimodality cardiovascular imaging of patients with pericardial disease: Endorsed by the society for cardiovascular magnetic resonance and society of cardiovascular computed tomography. J Am Soc Echocardiogr [Internet]. 2013;26(9):965–1012.e15. Available from: http://dx.doi.org/10.1016/j.echo.2013.06.023

10. Troughton RW, Asher CR, Klein AL. Pericarditis. 2004;363:717-27.

11. Mikolich JR. New Diagnostic Criteria for Acute Pericarditis: A Cardiac MRI Perspective. 2015;15:1-7.

12. Grove WM, Zald DH, Lebow BS, Snitz BE, Nelson C. Clinical versus mechanical prediction: A meta-analysis. Psychol Assess. 2000;12(1):19–30.

13. Yan AT, Yan RT, Tan M, Casanova A, Labinaz M, Sridhar K, et al. Risk scores for risk stratification in acute coronary syndromes: Useful but simpler is not necessarily better. Eur Heart J. 2007;28(9):1072–8.

14. Denmark KT, Bax JJ, Morrow DA, Task A, Members F, Kristian C, et al. Fourth universal definition of myocardial infarction (2018). 2018;(August):1-33.

15. Higgins, Charles B; Finck S. Constrictive Pericarditis and Restrictive Cardiomyopathy: Evaluation with MR Imaging. Radiology. 1992;

